# Supportive Mobility Across the Lifespan in Cerebral Palsy: A Modified Delphi Study to Establish Stakeholder Research Priorities

**DOI:** 10.1101/2022.01.26.22269919

**Authors:** Heather A. Feldner, Deborah Gaebler-Spira, Varun Awasthi, Kristie Bjornson

## Abstract

**Aim:** The overarching aim of this research was to 1) Understand the mobility experiences, supported mobility device (SMD) use, and desired participation outcomes of people with cerebral palsy (CP) across the lifespan; and 2) Describe how perspectives of rehabilitation care and professional resources may influence mobility decision-making processes and outcomes. The aim of this study was to co-develop research priorities and identify meaningful research questions with a diverse group of stakeholders representing the CP community for implementation in subsequent research activities.

**Methods:** A modified, three-round Delphi consensus study was conducted with a stakeholder advisory panel consisting of three adults with CP, two parents of children with CP, and four SMD providers.

**Results:** The advisory panel identified 13 unique topical categories focused on SMD selection and use, stratified by age group and stakeholder role. Questions or statements within each category were ranked, and top consensus and concordance statements were retained, reviewed, and refined for use in a co-developed focus group guide.

**Interpretation:** A modified Delphi process was a useful tool for stakeholders in co-developing research priorities related to SMD use across the lifespan. Drawing on the lived expertise of stakeholders is important in facilitating improved research translation in the CP community.

**What this Paper Adds:**

- Nine stakeholders from the CP community participated as Stakeholder Advisory Panelists and co-developers of research tools
- Stakeholders identified 175 unique responses across 12 SMD related categories
- Stakeholders prioritized 38 mobility technology research priorities during consensus-building
- Results from consensus-building will be directly implemented into a qualitative focus group protocol

---

Cerebral Palsy (CP) is the most common childhood onset motor disability, and in the US today, nearly 800,000 people are living with CP.^1^ While each individual living with CP is unique, key rehabilitation outcomes across the lifespan focus on acquisition and maintenance of mobility, whether through walking independently or through the use of supportive mobility devices (SMD) such as walkers, wheelchairs, or crutches.^2,3^ SMD is important for people with CP at any age but especially as motor ability changes over time.^4^ Many factors such as pain, body mass changes, and fatigue impact can impact mobility and walking ability across the lifespan, and over half of children with CP who are initially ambulatory experience regression in walking ability by adulthood.^5,6^ From orthotics to complex powered mobility devices, SMD has been shown to improve function, activity, and participation.^4,7-9^ However, issues of social stigma, device design/size, cost, and lack of accessibility in the built environment remain, presenting significant barriers to individuals with CP who use SMD.^8,10-12^

Existing literature describes these access barriers, along with mobility trends over time, biomechanical aspects of SMD use, perceptions of users and caregivers regarding their SMD, and decision-making guidelines for consideration of certain types of mobility technology, such as features of walkers or gait trainers, or introducing powered mobility when children may not walk, lose the ability to walk efficiently, or be significantly delayed in walking.^13,14^ In a study that examined perceived areas of importance for therapeutic intervention, youth with CP expressed that SMD was a priority, even when caregivers and healthcare providers expressed different priorities.^15^ Research also offers a snapshot of the realities of SMD provision in various settings, reporting that many clinicians lack training altogether, or lack confidence in their training and ability to evaluate, recommend, and adjust SMD for their clients.^16,17^ Despite this concerning gap, the literature underscores the importance of appropriate SMD, matched with appropriate and accessible environments, in maximizing mobility and participation outcomes.^7-9^

For children and adults with CP, SMD is a ubiquitous part of the lived experience, and thus an area of priority for further development and process improvement. However, little of this work has been co-developed by stakeholders from within the CP community or has involved shared-decision making recommendations specifically develop to support the procurement and use of SMD. Two examples from the current literature are notable exceptions. First, inclusion of people with CP specifically in the process of developing a focused research agenda was documented in Australia employing a virtual Delphi process.^18^ Additionally, in 2017 a group of nearly 50 stakeholders including individuals with CP, caregivers of children with CP, and clinicians convened at *Research CP*, a grant-funded conference to address the future of patient-centered CP research.^19^ The experiences and priorities that were shared specifically pointed to a gap in understanding and engaging in shared decision-making about SMD needs across the lifespan to minimize pain and fatigue and maximize activity, participation, and health in people with CP across all ages and functional levels.^19^ It is this work that has catalyzed the current study.

In recognizing stakeholder groups as experts in their field or in their lived experience, the Delphi method can be a useful research tool to employ. The Delphi method is widely used to understand and rank priorities from within a selected group of experts in a particular topic area, with an underlying assumption that group input is superior to individual input when establishing topical consensus.^20^ Delphi studies utilize multiple survey rounds to elicit open responses from participants, react and reply to responses of other group members, and modify or strengthen their own responses as a result of this reciprocal engagement. Although widely used across healthcare and other disciplines, the Delphi technique has only been minimally used in SMD research to date.^18,21^ Advantages of this technique include participant anonymity, which is intended to facilitate more open dialogue and support multiple opinions as consensus is sought, as well as the ability for the researcher to direct a controlled dialogue and feedback process toward an intended outcome.^20^ Anonymity between participants may be especially relevant in a mixed group of stakeholders, since power dynamics in healthcare and rehabilitation practice have historically favored healthcare professionals over people with lived experience of a health condition.^22^

This study represents the first stage of a multi-phase, mixed-methods research program that aimed to investigate the processes and outcomes of SMD procurement and use across the lifespan in adults and children with CP. Our overarching objectives were to 1) Understand the mobility experiences, supported mobility device (SMD) use, and desired participation outcomes of children, youth, and adults with CP; and 2) Describe how perspectives of rehabilitation care providers (therapists, physicians, durable medical equipment vendors) and professional resources (time, knowledge, etc.) may influence mobility decision-making processes and outcomes in people with CP and their caregivers. This paper describes the first study stage, a modified Delphi consensus study employed with a stakeholder advisory panel representing the CP community to leverage shared expertise in the co-development of study priorities and focused topical questions to be implemented in the subsequent phases of the research protocol.

## Method

This study was conducted with institutional approval from the [*institution removed for review]* Institutional Review Board (#1490). Prior to participating in study activities, participants provided written consent for all research procedures. The study was carried out by a research team consisting of a rehabilitation physician and two pediatric physical therapists, all with extensive professional experience in working with SMD and partnering with individuals with CP of all ages and functional levels.

### Participants

Nine participants were recruited from within and around the CP community for a stakeholder advisory board. Purposive sampling was conducted to ensure representation from three stakeholder groups: 1) Individuals with CP; 2) Caregivers of individuals with CP; and 3) Healthcare professionals with expertise in SMD actively serving clients with CP. The advisory board consisted of three adults with CP, two parents of children with CP, and four SMD providers, who represented fields of Occupational and Physical Therapy and SMD manufacturing. Two of the adults with CP also had careers as physicians, thus offering multiple lived perspectives. Eight of the panelists were white, one was Asian, and one of the parents was the adoptive mother of an African American child. Panelists represented geographic regions across the US, including the Pacific Northwest, Midwest, South, and East. Each panelist was paid a $500 honorarium for between 12-15 hours of their time across a year-long period. In addition to completing the Delphi consensus process, panelists assisted in data analysis in subsequent study stages.

### Study Procedures

A modified Delphi consensus technique was used to develop prioritized and meaningful content for use within subsequent research procedures, specifically topic content and questions for a qualitative focus group guide. Participants engaged in three rounds of online surveys, consistent with recommendations in previous literature.^23^ A schematic of the full Delphi process is included in Figure 1.

**Figure 1.**
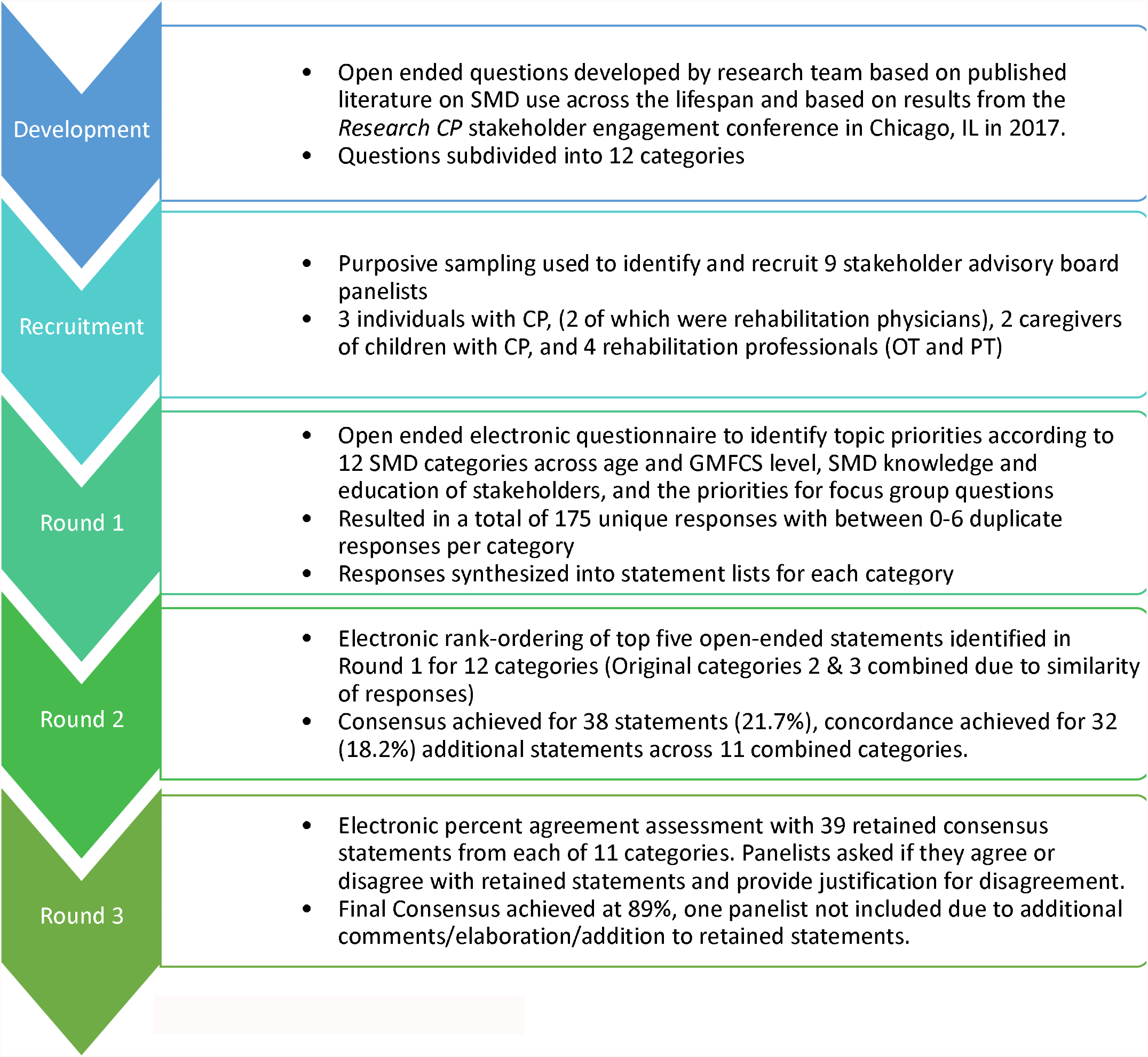
Delphi Consensus Building Process. This modified Delphi study employed three rounds of consensus building to determine topical priorities and questions for a focus group guide to be used in a subsequent qualitative study. An open-ended first round resulted in statements from panelists across 12 categories. The second round involved rank ordering the top five statements in 11 categories in order of importance (two categories were combined in Round 2 due to the similarity of responses in Round 1), and the third round presented the rank-order results and assessed agreement between stakeholder panelists.

#### Delphi Round One: Open Ended Questionnaire

A first round electronic survey presented a series of open-ended questions to gather responses regarding the most important considerations for SMD selection and use across the lifespan for people with CP. Open-ended questions were stratified into categories that considered age, functional level, and stakeholder role. A full listing of Round One categories is presented in Table 1.

**Table 1.** Delphi Round 1 Open Ended Question Categories

| Category | Question |
| --- | --- |
| 1 | Most Important Considerations for young children regarding SMD selection and use |
| 2* | Most important considerations for adolescents regarding SMD selection and use |
| 3* | Most important considerations for young adults regarding SMD selection and use |
| 4 | Most important considerations for older adults regarding SMD selection and use |
| 5 | Most important considerations for individuals with CP in GMFCS levels 2 & 3 |
| 6 | Most important considerations for individuals with CP in GMFCS levels 4 & 5 |
| 7 | Most important for suppliers and manufacturers of SMD to know |
| 8 | Most important for physicians and therapists to know about SMD |
| 9 | Most important for individuals with CP and their families to know about their options for SMD |
| 10 | Most important focus group questions to ask individuals with CP and their families |
| 11 | Most important focus group questions to ask physicians and therapists |
| 12 | Top priorities for SMD design and provision for individuals with CP over the next decade |

\*Categories 2 and 3 were combined following Round 1 due to the similarities in panelist responses.

#### Delphi Round Two: Ranking Responses

In the second round, an electronic survey provided an aggregate, anonymous summary of responses from Round One. Responses were provided in no particular order, though duplicate responses across panelists were denoted with asterisks to indicate a high-frequency response. Within each category, panelists were asked to rank-order the top five most important topic statements or questions they wished to see carried forward in the focus group guide. Panelists were also given an opportunity to provide free responses for each category asking what (if anything) should be added, removed, or refined from the list. See Figure 2 for an example of the Round Two survey.

**Figure 1.**
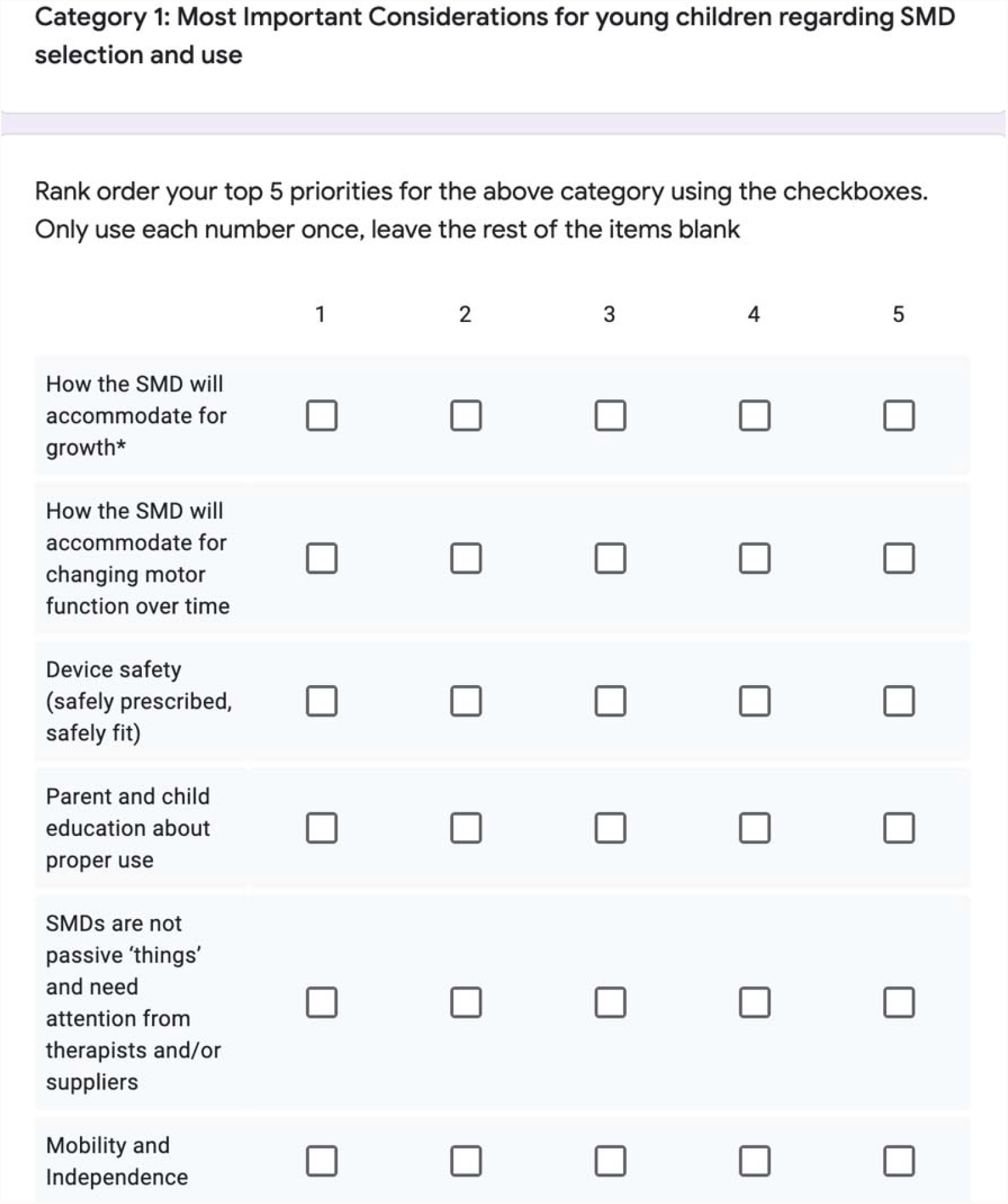
Delphi Round 2 Rank Ordering Sample. An example of the Round 2 Delphi rank ordering exercise for Category 1 from Round 1. Panelists were asked to rank order the top five most important statements within each category and leave the remaining statements in each category blank.

#### Delphi Round Three: Rankings and Agreement

The final round electronic survey provided the panel with the ranking results tallied from Round Two. Panelists were instructed to indicate whether they agreed or disagreed with the rank ordering. If disagreement was indicated, panelists were asked to provide rationale about why they remained outside of consensus. Feedback from this final round was used to structure focus group topics and prioritize targeted questions for the qualitative focus group guide.

### Data Analysis

Content analysis of open-ended Round One results was conducted to understand stakeholder priorities across categories and consolidate like responses into statements presented in Round Two. Quantitative study results from Round Two were analyzed using descriptive statistics (μ, SD) to determine the average ranking and dispersal of statements retained in each category. Due to the small sample size, consensus for statement retention was pre-defined as >/=50% respondent agreement prior to analysis. If between 40-50% agreement was reached, these statements were labeled as concordant, to denote agreement that did not meet the threshold for consensus. Concordant statements were retained for review by the research team, but were not presented to panelists during Round Three. All statements below 40% agreement were discarded. In Round Three, percent agreement was calculated for the retained statements and any open-ended justification of disagreement comments were analyzed qualitatively for content.

## Results

Results from Round One generated 175 distinct responses across 12 categories. Each category had at least one duplicate response across the panel members, with the exception of category 11 (*Priorities for clinician focus group questions*). Categories with the most duplicate responses included categories 2 and 3 (*Most important considerations for adolescents and young adults regarding SMD*) and category 12 (*Priorities for SMD design over the next decade*), with six responses in each category reported by multiple panelists. Because responses in categories 2 and 3 were so similar, these were combined during subsequent Delphi rounds. Categories with the least crossover in response included category 4 (*Considerations for older adults regarding SMD*), category 8 (*Most important for physicians and therapists to know regarding SMD*), category 10 (*Priorities for focus group questions with individuals with CP and families*), with only one item in each category reported by multiple panelists, indicating a wider dispersal of responses.

Round Two results were mixed. Rank ordering of statements resulted in clear consensus and retention of 38 statements (21.7%), with at least one retained statement across each of the 11 consolidated categories (μ=3.45, Range 1-6 per category). There were 32 additional statements identified as concordant (18.2%), with a lower mean but similar range across categories (μ=2.90, Range 1-6 per category). Categories with the greatest number of consensus statements retained included *Most important considerations for young children regarding SMD* (Category 1), *Most important considerations for individuals with CP in GMFCS levels 2 & 3* (Category 5), and *Most important for physicians and therapists to know about SMD* (Category 8). Categories with the most variability in ranking, and thus the fewest number of consensus statements retained, included *Most important focus group questions to ask individuals with CP and their families* (Category 10), *Most important focus group questions to ask physicians and therapists* (Category 11), and *Top priorities for SMD design and provision for individuals with CP over the next decade* (Category 12). Notably, both Categories 10 and 12 had correspondingly high concordance frequencies, with 6 and 5 statements reaching concordance, respectively. A full listing of the consensus and concordance statements retained for each category are listed in Table 2.

**Table 2.**
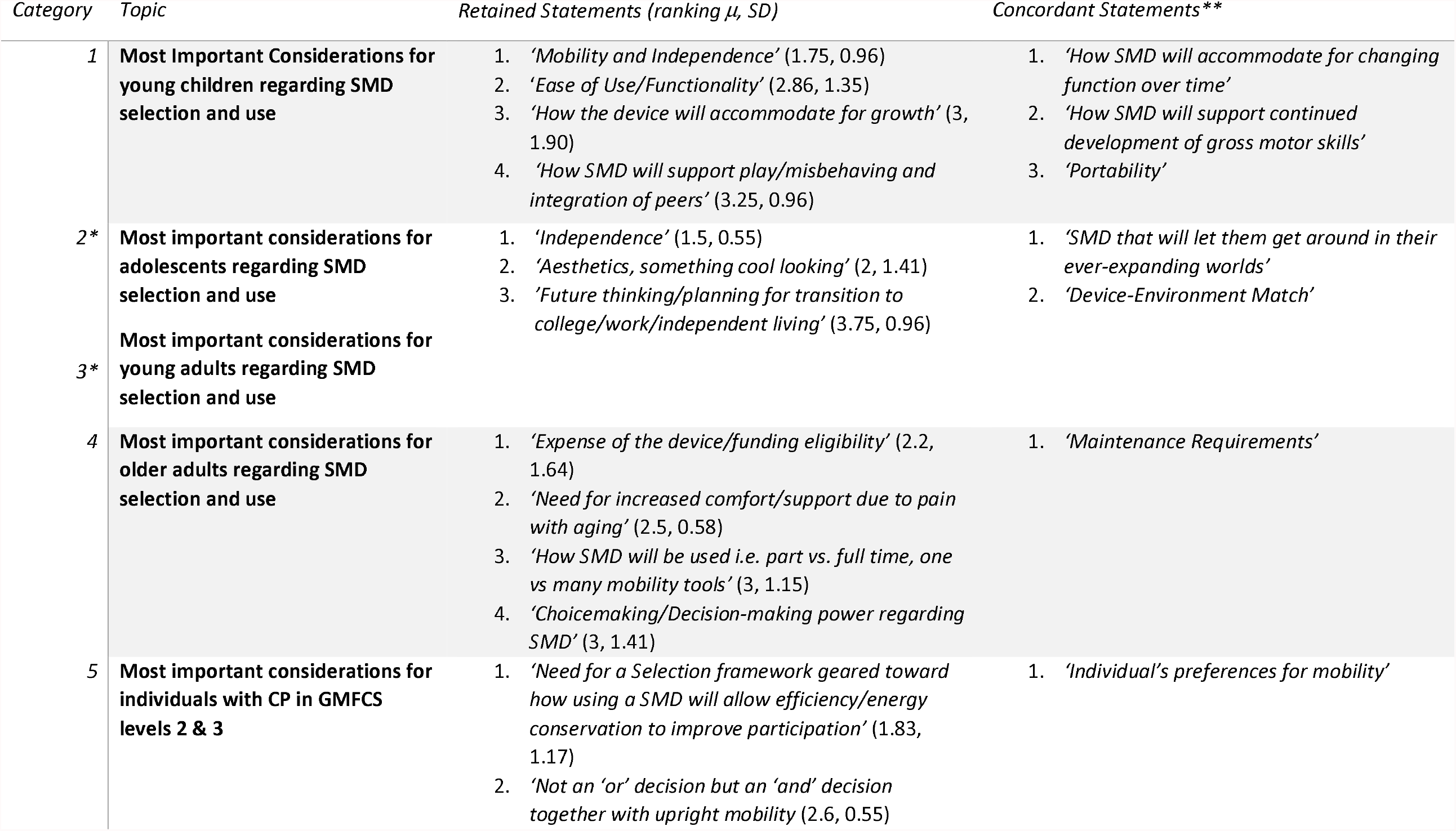
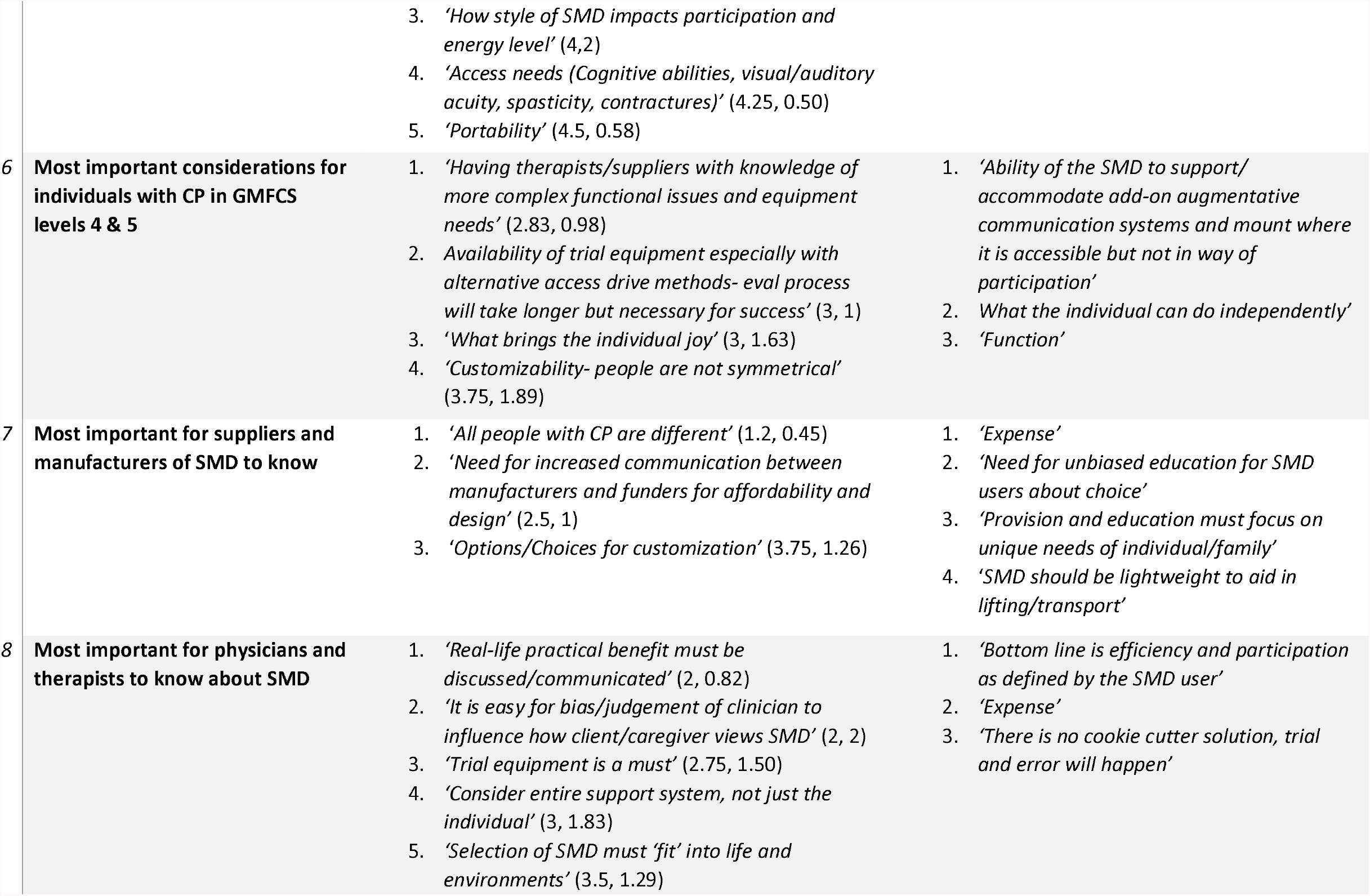
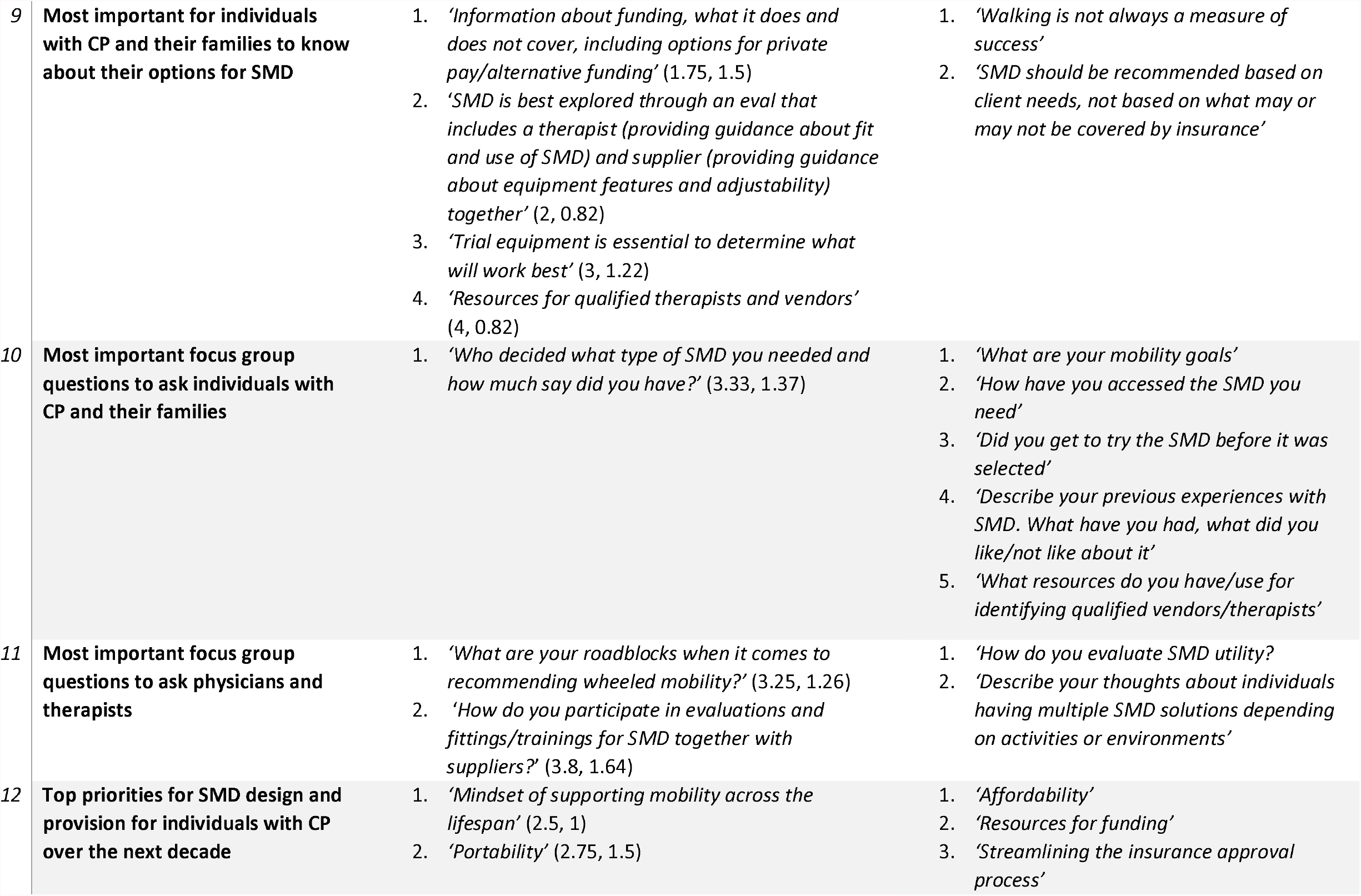
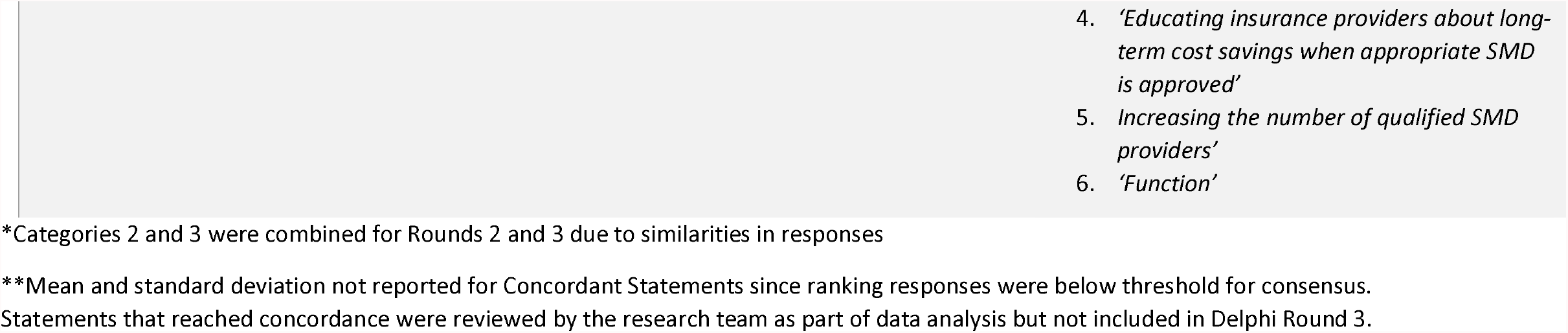
Rank Ordered Results from Delphi Round 2 Per Category

Open-ended feedback from panelists highlighted additional areas of importance, including clinician continuing education related to SMD, identification of policy barriers and resources for funding, acceptability of SMD for participation in desired activities, and the desire to preserve physical status and energy with a multimodal (‘And’ rather than ‘Or’) approach to SMD.

Round three results indicated that panelists reported 89% agreement (n=8) with the aggregate ranking results and retained statements. One panelist provided additional feedback for several categories, this feedback was discussed by the research team and incorporated into the subsequently developed research materials, however, the research team did not feel this feedback warranted a subsequent Delphi round since overall consensus was high and the panelist’s open-ended responses stated baseline agreement but expanded the discussion, rather than representing a fundamental disagreement.

## Discussion

This study, the first step in a mixed-methods, multi-phase research project, convened a stakeholder advisory panel from the CP community to understand important considerations and set priority topic areas and questions for subsequent research activities. A modified Delphi technique was used to establish consensus across 12 categories related to SMD, and to inform the subsequent conduction of the next study phase.^20^

Consensus (>/= 50%) was met for 38 statements across a combined 12 SMD related categories. The data also showed *stability* (i.e. consistency across categories without necessarily reaching pre-defined levels of consensus).^24^ This moderate rate of consensus was surprising at first but may be explained by the distribution of unique stakeholder experiences and perspectives within the panel. Because the survey responses were anonymous, however, it is impossible to determine if panelist characteristics played a role in ranking responses. Nevertheless, this moderate rate of clear consensus is mirrored in other recent Delphi studies involving CP research, and did not appear to affect the high final round agreement toward the top-ranked statements.^23^

Many similarities across categories emerged, highlighting the salience of the consensus and concordance statements in regard to stakeholder priorities for research. For example, statements about the importance of trial equipment were ranked among the top three statements in two related categories (*Most important for individuals with CP and their families to know about SMD* and *Most important for physicians and therapists to know about SMD)*. This is consistent with extant literature describing barriers and facilitators to SMD use, including the presence or lack of trial equipment, by many individuals with disabilities and their caregivers.^10,25^ Similarly, across two age-group related categories (*Most important SMD considerations for Adolescents* and *Young Adults)*, panelists noted priorities related to device aesthetics, as well as future-oriented thinking and planning for transition to college, work, or independent living, which is also consistent with literature describing the importance of transition services and education as young people with CP assume more responsibility for their healthcare and participation.^26,27^

Across multiple categories, provider knowledge and qualifications related to SMD emerged as a priority. This was salient from the perspective of empowering individuals with CP to seek out knowledgeable healthcare professionals as well as providers themselves taking responsibility for their own learning about SMD as a topic not largely covered during professional training for physicians and therapists. Numerous studies report the need for additional competency related to assistive technology provision and policy.^16,17^ What remains unclear is whether individuals with CP and their families understand what to look for regarding provider credentials or experience, or whether existing clinicians understand how and where to access additional training resources in increasingly busy practice environments.

Finally, panelist rankings for all age and GMFCS group categories as well as priority focus group questions for individuals with CP indicated a desire to focus on activity and participation-level domains as well as shared decision making related to SMD in future research. This also mirrors previous recommendations for the deployment of user-centered frameworks that placed corresponding emphasis on the person or user and their desired mobility goals, as well as the healthcare providers or clinical setting to ensure well-matched technology provision processes and products.^7,8^ Panelists felt this was a central issue to discuss with focus group participants, as it remains unknown how effective these methods of person-centered care for shared decision-making are played out in every day SMD provision.

### Study Limitations and Future Directions

This study has several limitations. First, though the research team has professional experience in working with SMD and individuals with CP across the lifespan, it is important to note that no one has lived experience of CP. For this reason, it was important to directly involve individuals with CP in our stakeholder advisory panel to ensure that the needs and priorities of the CP community were represented by those with lived experience of SMD use.^7,28^ Additionally, the stakeholder advisory panel was comprised of only nine members, and though response rate was generally good, not all members took part in every Delphi round, thus limiting the generalizability of our results. However, current literature indicates that successful Delphi studies in healthcare may involve anywhere between 6-30 individuals, therefore the advisory panel group size remained in acceptable range.^23,29,30^ Further, all advisory panelists were from the US, so responses and priorities were limited to experiences within the US SMD provision system.

Future research should continue exploring stakeholder-led activities to develop meaningful research priorities and questions for the CP community, and include funding agencies as key stakeholders. While this grassroots involvement has occurred in small pockets, opportunities exist for broader-scale agenda development.^19^ Academic and clinical researchers and healthcare providers can leverage CP community networks to further successful translation of research into direct benefit for people with CP across the lifespan.

## Conclusion

This study was the first phase of an overarching study to understand the mobility experiences, supported mobility device (SMD) use, and desired participation outcomes of children, youth, and adults with CP. Consensus was established for numerous statements related to research priorities and topic areas for SMD provision and use across the lifespan, which were subsequently employed in additional research activities. A Delphi process with stakeholders in a given community of study appears to be a valuable method to ensure the setting of a meaningful agenda in CP research.

## Supporting information

STROBE Checklist

## Data Availability

All data produced in the present study are available upon reasonable request to the authors.

## List of Abbreviations

SMD: Supportive Mobility Device
CP: Cerebral Palsy

## Acknowledgements

The authors would like to thank the members of the stakeholder advisory panel that contributed to making this project successful, and the Cerebral Palsy Research Network for their feedback in developing the overarching project.

